# Smartphone Movement Sensors for Remote Monitoring of Respiratory Rate: Observational Study

**DOI:** 10.1101/2021.12.03.21267247

**Authors:** Sophie Valentine, Benjamin Klasmer, Mohammad Dabbah, Marko Balabanovic, David Plans

## Abstract

**Background:** Mobile health offers potential benefits to patients and healthcare systems alike. Existing remote technologies to measure respiratory rate (RR) have limitations, such as cost, accessibility and reliability. Using smartphone sensors to measure RR may offer a potential solution.

**Objective:** The aim of this study was to conduct a comprehensive assessment of a novel mHealth smartphone application designed to measure RR using movement sensors.

**Methods:** In Study 1, 15 participants simultaneously measured their RR with the app, and an FDA cleared reference device. A novel reference method to allow the app to be evaluated ‘in the wild’ was also developed. In Study 2, 165 participants measured their RR using the app, and these measures were compared to the novel reference. Usability of the app was also assessed in both studies.

**Results:** The app, when compared to the FDA-cleared and novel references, respectively, showed a mean absolute error (MAE) of 1.65 (SD=1.49) and 1.14 (1.44), relative MAE of 12.2 (9.23) and 9.5 (18.70) and bias of 0.81 (limits of agreement (LoA) =-3.27-4.89) and 0.08 (−3.68-3.51). Pearson correlation coefficients were 0.700 and 0.885. 93% of participants successfully operated the app on their first use.

**Conclusions:** The accuracy and usability of the app demonstrated here show promise for the use of mHealth solutions employing smartphone sensors to remotely monitor RR. Further research should validate the benefits that this technology may offer patients and healthcare systems.

## Introduction

Extensive growth in the development and adoption of remote healthcare tools has been seen in recent years in response to increasing demand for traditional offerings. Notably, the COVID-19 global health pandemic has made salient how these mobile health (mHealth) tools may support healthcare systems to manage their patients when resources are pushed to breaking point.^1-3^ As more widely accessible tools can be used by more people - and therefore offer greater impact - many mHealth smartphone applications (apps) have been developed, due to the high global penetration of smartphones. These systems offer a wide variety of services from telemedicine to remote monitoring and self-care, and evidence suggests they may produce improved economic^4^ and health outcomes.^5^

Respiratory rate (RR) is a fundamental indicator of health status for many health conditions, both general and specific to the respiratory system.^6-11^ As such, mHealth solutions for monitoring of RR may offer significant value to patients and healthcare professionals (HCPs) alike. Although several such solutions exist, many fall short on various factors. Hardware-based solutions, including piezoelectric sensors^12^ pulse oximeters,^13^ and multi-sensor devices,^14-15^ are typically expensive, vulnerable to limited means of manufacture and distribution,^16^ and may lack interoperability with other health records, which is cited as a critical risk to decentralisation of national healthcare systems.^17^ Software-based solutions address limitations of cost, manufacture and distribution; however, they typically employ less-stable mechanisms of action. These mHealth apps often use smartphone cameras or microphones,^18-20^ the latter of which have been evidenced to be vulnerable to environmental noise at the cost of accuracy and usability.^21-22^

Movement sensors may present a promising alternative software-based solution for mHealth RR monitoring. Research indicates that multi-axial accelerometers and gyroscopes - as found ubiquitously in modern smartphones - can accurately capture RR based on chest movements.^23-30^ Additionally, due to their mechanism of action, these sensors are significantly less affected by environmental noise. Overall, smartphone-based measurement of RR provides a potential low cost, and widely available method for RR measurement, both in a remote monitoring environment, or in locations where specialised hardware and software are not available.

This article presents an observational assessment of a novel user-centric mHealth smartphone app that measures RR using smartphone movement sensors. We first conducted a preliminary evaluation of the device and study methods via a small lab-based study, then jointly assessed accuracy and usability on a greater scale and ecological valid environment via a remote study. Ethical approval was provided by the University of Exeter’s Research Ethics Board (application ID eUEBS004088) and all research was conducted in compliance with the Declaration of Helsinki.

## Study 1

### Methods

The preliminary evaluation pursued three aims: (1) to establish the accuracy of the novel mHealth smartphone app relative to a reference device cleared by the US Food and Drug Administration (FDA), (2) to understand the usability of the mHealth app and (3) to evaluate the suitability of a novel reference method that would permit accuracy assessments to be conducted via remote and real-world studies. Through a prospective, non-interventional, non-randomised study conducted on healthy volunteers, RR estimates provided by the FDA-cleared reference device were compared to those from the novel mHealth smartphone app and the novel reference.

#### Measurements

##### Novel mHealth smartphone app

The mHealth app contained a purpose-built user interface (Figure 1). The user is instructed to hold their smartphone to their upper-middle chest with the screen facing outwards while sitting still and breathing normally for the duration of the 30-second sensor recording. Data is captured from the smartphone’s tri-axial gyroscope and interpolated to achieve an even 100Hz sample frequency. A low-pass Butterworth filter with 0.4 Hz cut-off is applied to remove high-frequency noise while retaining activity associated with breathing, typically in the 0.16-0.33Hz range (10-20 breaths per minute (BPM)). RR is calculated by performing an autocorrelation before normalising the resulting signal. A peak-finding routine then identifies prominent peaks corresponding to the cyclical property of breathing movements. The mean inter-peak interval (IPI) is then calculated and converted to a ‘per minute’ RR estimation by division by 60 (seconds) (Figure 2).

**Figure 1.**
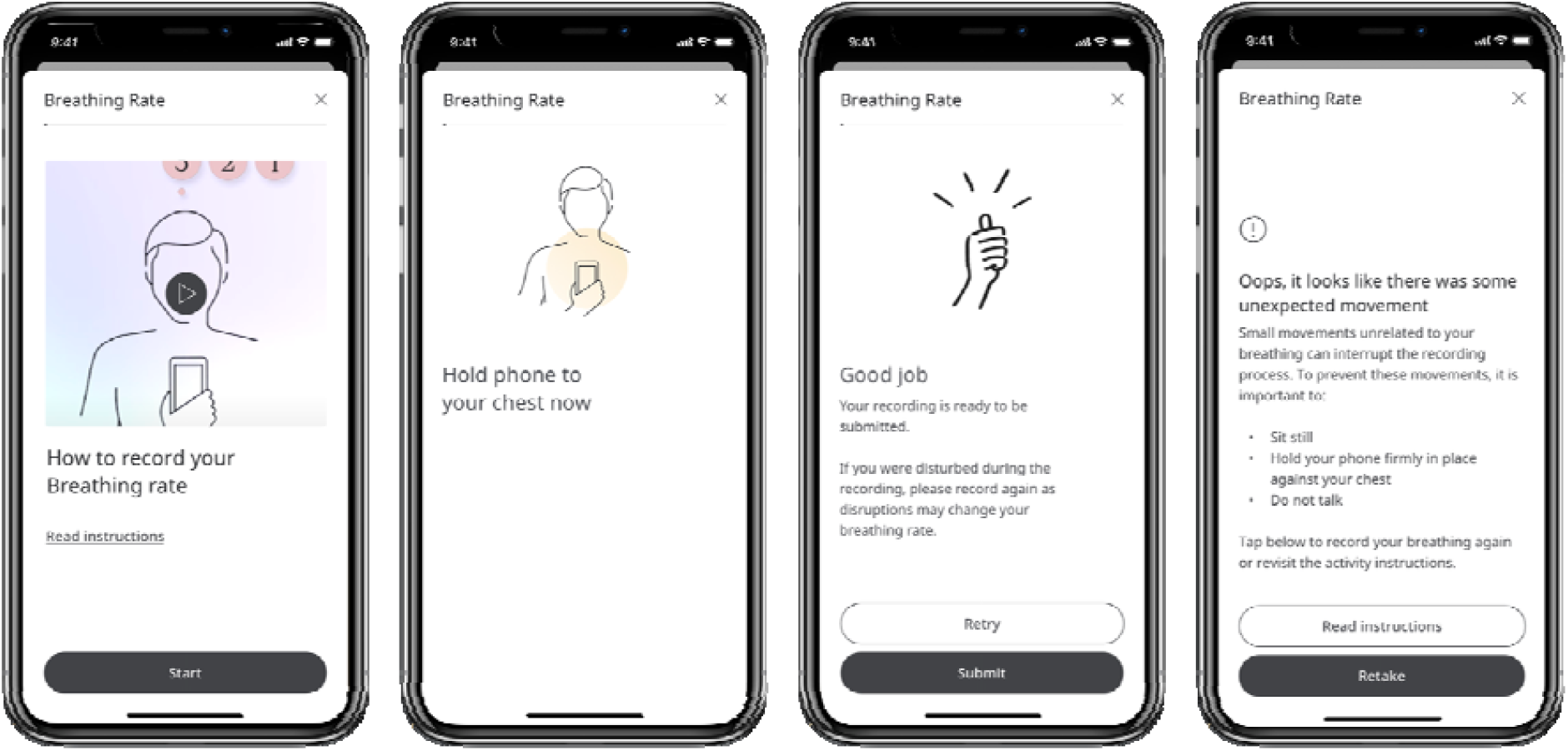
Selected wireframes from the user interface of the mHealth app, depicting (from left to right) an option given to the user to view operation instructions in video or written format, an instruction for the user to hold their smartphone to their chest, a clinical safety feature allowing the user to retake a recording if they were disturbed while taking the original recording, and feedback given to the user if their recording fails the signal check.

**Figure 2.**
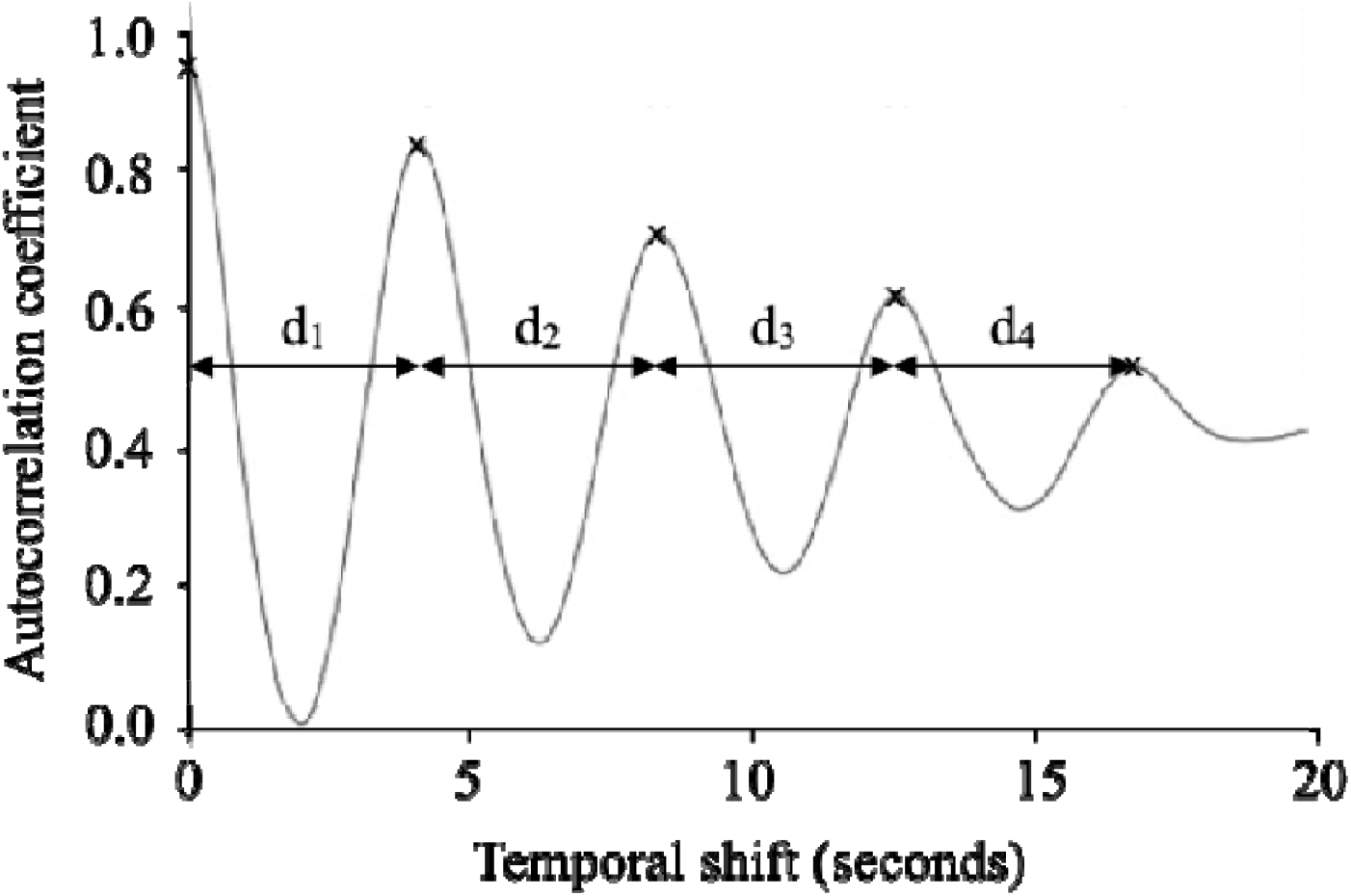
Graphical depiction of peak finding following application of an autocorrelation method. The grey line shows an example of correlation coefficients for a movement sensor (gyroscope) signal correlated with itself at progressive temporal shifts. Black crosses indicate prominent peaks corresponding to the cyclical property of breathing movements. IPIs are depicted by d_i_.

An additional ‘signal check’ routine assesses whether the signal quality is sufficient to accurately derive RR, based on whether the number of autocorrelation peaks or standard deviation (SD) of the individual IPIs meet predetermined thresholds identified via preliminary bench-testing. If a recording fails the signal check, the user is informed via the app’s UI, redirected to the operation instructions and prompted to try again. Passing the signal check within three recordings attempts constitutes a successful use of the system, and three consecutive signal check failures constitute an unsuccessful use of the system, after which the user is instructed to seek support or try again later.

##### FDA-cleared reference

The *MightySat Rx*^13^, developed by Masimo Corporation, was selected as a reference due to its FDA-cleared status, continuous measurement and ease of use. The fingertip pulse oximeter derives RR using photoplethysmography (an optical measure of volumetric changes in peripheral blood flow). Continuous estimates of RR produced by this reference were converted to single weighted averages to facilitate comparison with data derived from the mHealth app.

##### Novel reference

The novel reference method involves identification of repeated cyclical peak-trough complexes within smartphone movement sensor signals(Figure 3). Signals of insufficient quality to derive RR are considered to fail the reference method. This method is conceptually similar to reference methods described in peer-reviewed literature reporting accuracy assessments of multiple RR devices, including successful FDA market clearance applications.^13; 31-33^ This method would permit accuracy assessments to be conducted via remote and real-world studies without a need for additional hardware, offering significant value in terms of research scale, cost and ecological validity via avoidance of observation bias.

**Figure 3.**
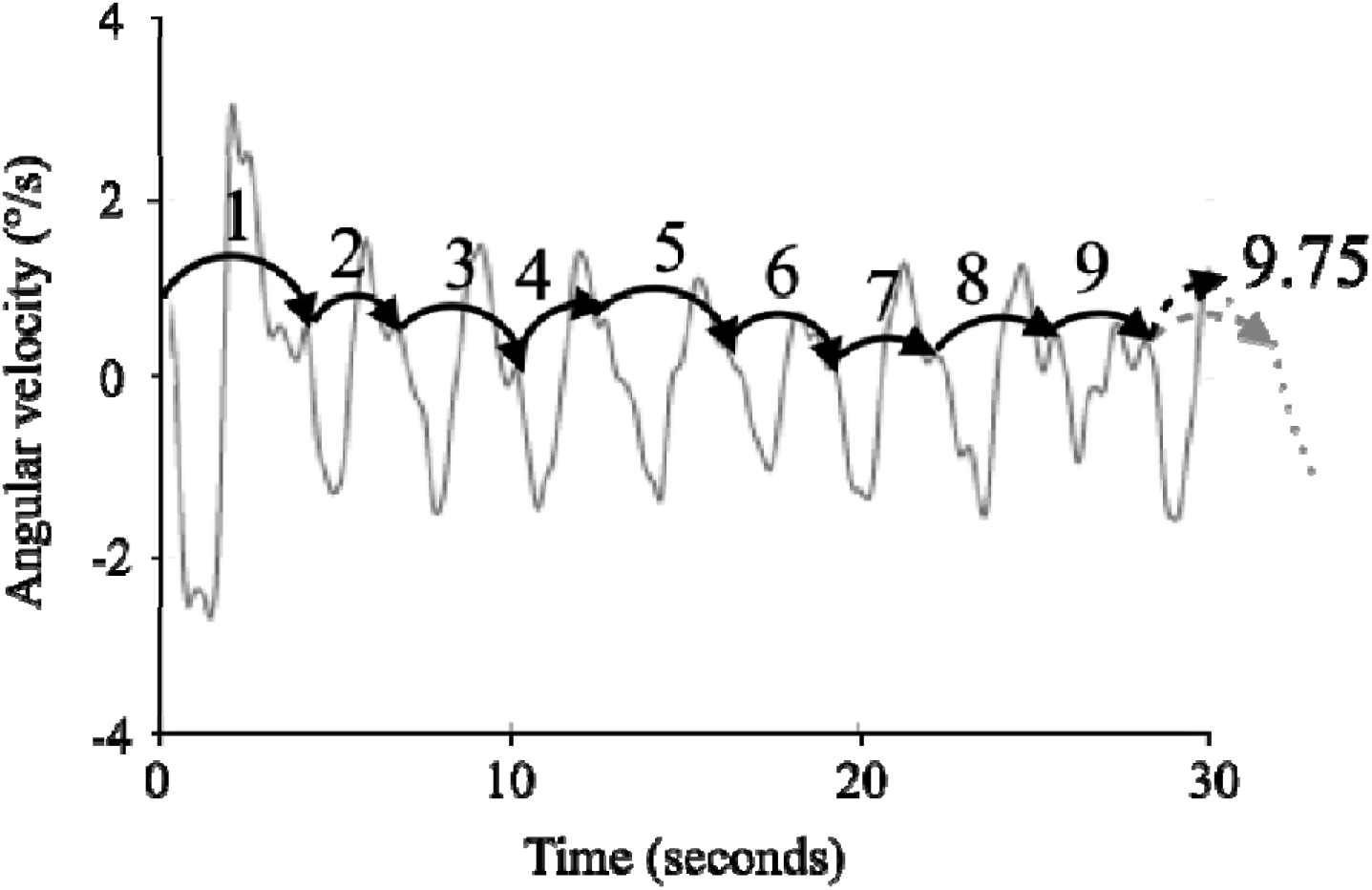
Graphical depiction of the novel reference method involving visual inspection of smartphone movement sensor signals by trained clinical and scientific researchers. The grey solid line shows an example of a smartphone movement sensor (gyroscope) signal, with black solid arrows depicting nine full repeated cyclical peak-trough complexes. The grey dotted line indicates the projected continuation of the movement sensor signal past the end of the recording period, with the grey dashed arrow indicating where a tenth full peak-trough complex would end. Hence, the movement sensor signal depicts a total of approximately 9.75 peak-trough complexes, with the final .75 peak-trough complex indicated by the black dashed arrow.

#### Participants and recruitment

Participants were recruited via convenience sampling. All were employees of the mHealth app manufacturer. Inclusion criteria included being aged 18 or over and being willing and able to follow the study protocol and complete an informed consent form.

#### Procedure

The study took place at the offices of the mHealth app manufacturer. Participants were provided with complete information concerning the study procedures and gave written informed consent to participate. The FDA-cleared reference device was applied to the forefinger of the participant’s left hand. Participants were provided with an iPhone XR with the mHealth app installed and received verbal instructions on operating the device: namely, to hold the smartphone to their upper middle chest with the screen facing outwards while sitting still and breathing normally during the 30-second recording. Participants were instructed to capture six recordings, disregarding whether each recording passed or failed the signal check. Audiovisual footage was captured during the study and used for offline synchronisation of data captured via the mHealth app and FDA-cleared reference. Specifically, this included sounds produced by the mHealth app indicating the start and end of the app’s recording period and depicting RR estimates displayed on the FDA-cleared reference’s monitor. Participation took around 10 minutes per participant.

#### Statistics

The error of the mHealth app and novel reference relative to the FDA-cleared reference was assessed through measures of mean absolute error (MAE), relative MAE and using the Bland-Altman method.^34^ Due to the non-normal distribution of absolute error data, confidence intervals for MAE and relative MAE were derived via bootstrapping with replacement employing 1000-iterations and a sample size of 100%. The proportion of clinically significant errors, defined as an absolute error greater than three breaths per minute,^35-36^ was also calculated. Direct relationships between RR estimates generated through the mHealth app, novel reference and FDA-cleared reference were assessed via Pearson Product Moment Correlation (PPMC). The usability of the mHealth app was assessed using the proportion and position of recordings that failed the signal check.

## Results

### Participants and data

15 participants took part in Study 1 (9 female), for whom 6 recordings each were collected for a total of 90. 26 (28%) mHealth app recordings failed the signal check and were excluded from analyses, resulting in a dataset of 64 paired samples. 29 (32%) of recordings failed the novel reference method, so were excluded from analyses, resulting in a dataset of 61 paired samples.

### Accuracy

#### mHealth app versus FDA-cleared reference

Error results indicated an MAE of 1.65 BPM (SD = 1.49) with a 95% confidence interval (CI) of 1.32-2.06. Relative MAE was 12.2% (SD = 9.23) with 95% CI of 10.06 - 14.57. Bias (FDA-cleared reference - mHealth app) was 0.81 (SD = 2.08) with limits of agreement (LoA) of -3.27 - 4.89, indicating RR underestimation by the mHealth app. 8 comparisons (12.5%) had an absolute error greater than 3 BPM. A Bland-Altman plot indicated error values as a function of RR averaged between the reference and mHealth app (Figure 4). *PPMC produced a coefficient of r(63) = 0*.*700, p < .000, indicating a high or strong association between the reference RR estimates and mHealth app RR estimate*^*37*^ *(Figure 5)*.

**Figure 4.**
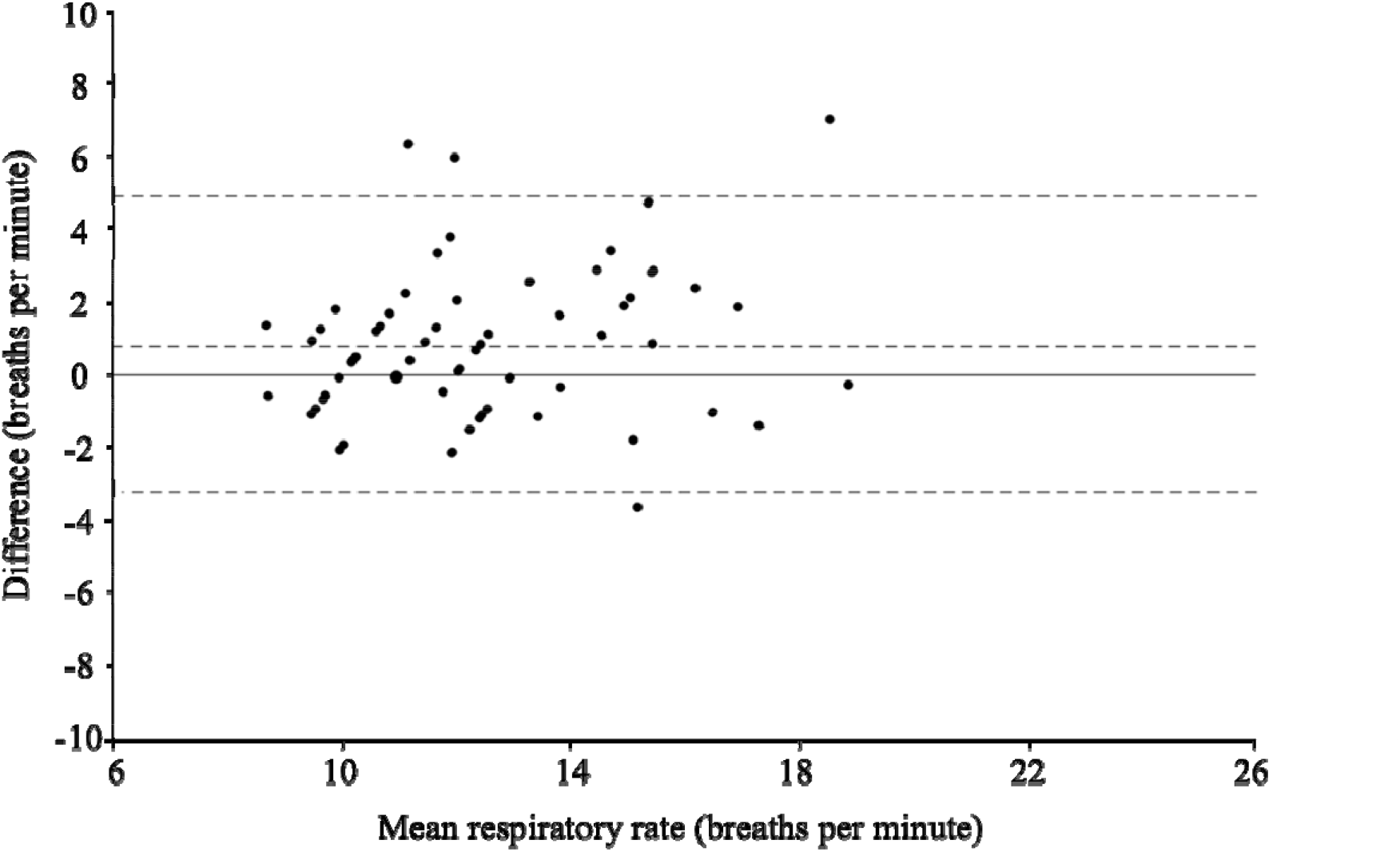
Bland-Altman plot for RR estimates provided by the mHealth app and FDA-cleared reference. The x-axis indicates RR estimates averaged between the mHealth app and FDA-cleared reference and the y-axis indicates the difference between RR estimates from each source (FDA-cleared reference - mHealth app). The solid horizontal line depicts a mean difference (bias) of 0 and dashed lines from top to bottom represent the upper limit of agreement (4.89), the observed mean difference (bias; 0.81), and the lower limit of agreement (−3.27). Marker size is proportional to the number of observations for each combination of values.

**Figure 5.**
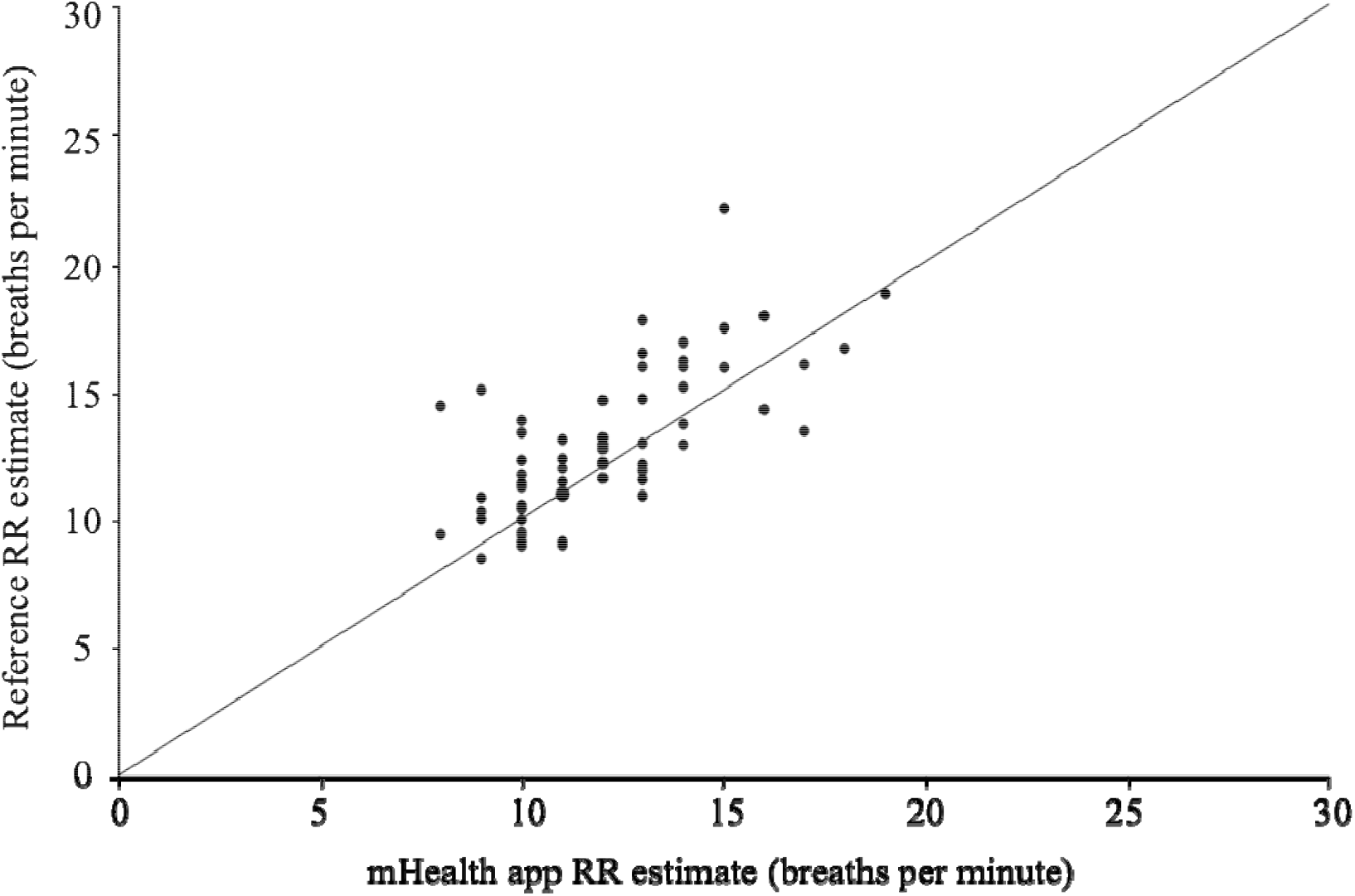
Scatterplot for simultaneous RR estimates provided by the mHealth app (x-axis) and FDA-cleared reference (y-axis). The solid line indicates the gradient y=x. Marker size is proportional to the number of observations for each combination of values.

#### Novel reference versus FDA-cleared reference

Error results indicated that MAE was 1.69 BPM (SD = 1.61) with a 95% CI of 1.23 - 2.22. Relative MAE was 12.8% (SD = 11.60) with 95% CI of 9.96 - 15.64. Bias (FDA-cleared reference - novel reference) was 0.22 (SD = 2.34) with LoA of -4.36 - 4.79, indicating slight RR underestimation by the mHealth app. 9 comparisons (15%) had an absolute error greater than 3 BPM. A Bland-Altman plot indicated error values as a function of RR averaged between the FDA-cleared and novel references (Figure 6). PPMC produced a coefficient of r(59) = 0.701, p < .000, indicating a high or strong association^37^ (Figure 7).

**Figure 6.**
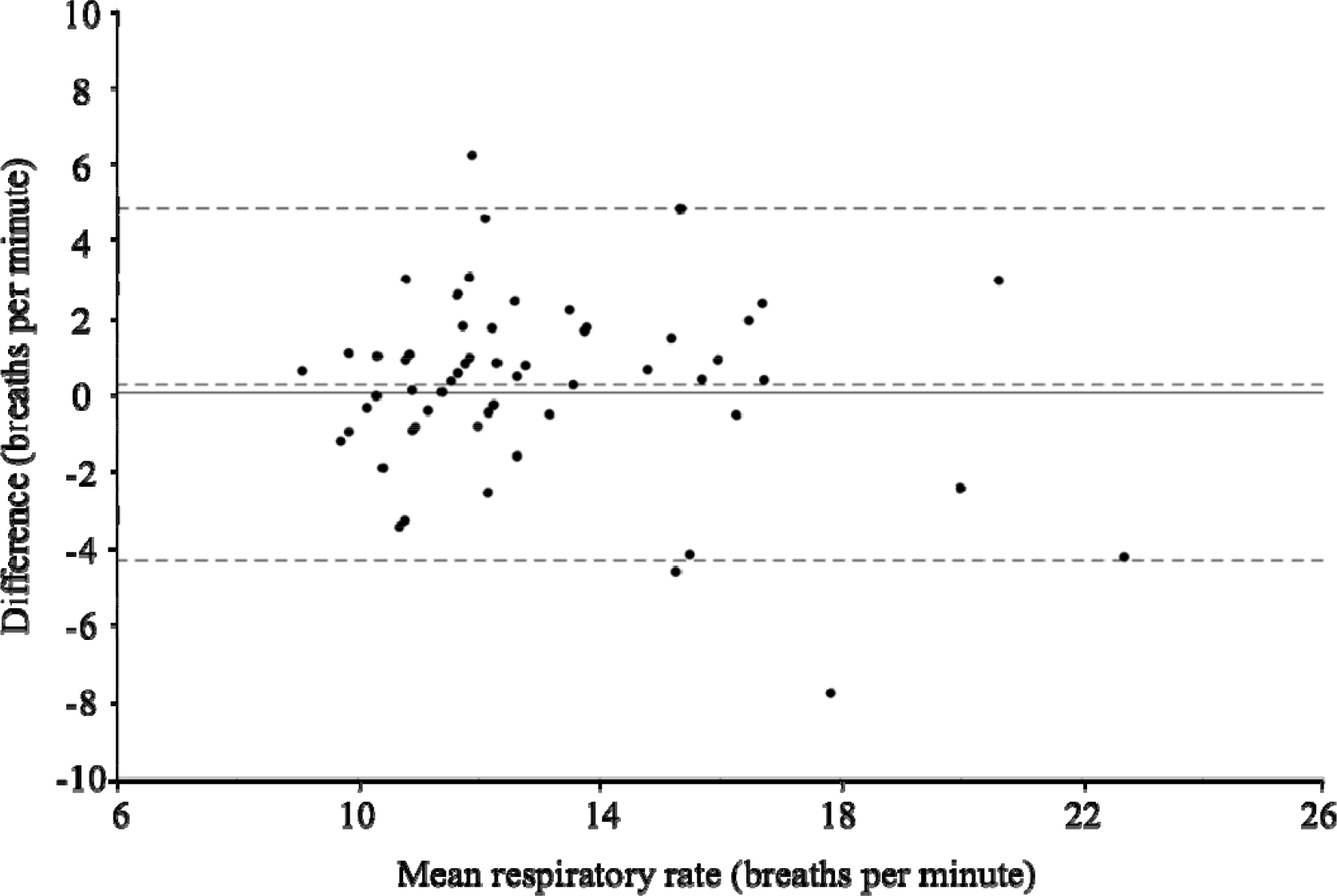
Bland-Altman plot for RR estimates provided by the novel reference and FDA-cleared reference. The x-axis indicates RR estimates averaged between the novel and FDA-cleared references and the y-axis indicates the difference between RR estimates from each source (FDA-cleared reference - novel reference). The solid horizontal line depicts a mean difference (bias) of 0 and dashed lines from top to bottom represent the upper limit of agreement (4.79), the observed mean difference (bias; 0.22), and the lower limit of agreement (−4.36). Marker size is proportional to the number of observations for each combination of values.

**Figure 7.**
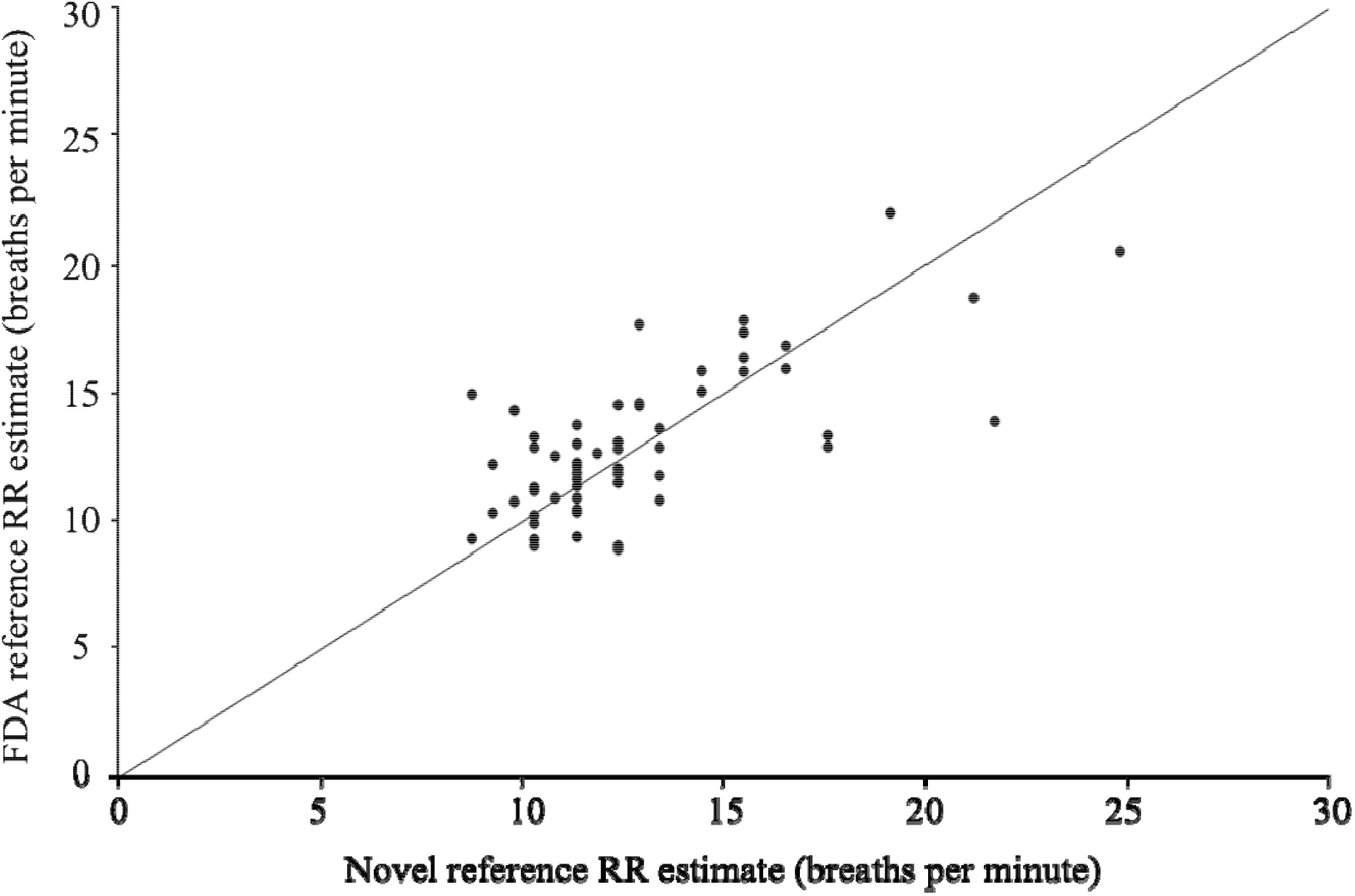
Scatterplot for simultaneous RR estimates provided by the novel reference (x-axis) and FDA-cleared reference (y-axis). The solid line indicates the gradient y=x. Marker size is proportional to the number of observations for each combination of values.

### Usability

14 of a total of 15 participants (93.3%) were able to use the system successfully on their first try (Table 1; Figure 8). Specifically, this indicates that they could capture one or more recordings that passed the signal check within the first three attempts. All participants were able to use the system successfully by the end of their second try.

**Table 1.** Number and proportion of participants able to use the system successfully on consecutive attempts in Study 1 (n = 15).

| Use of system, n (%) | Recording attempt |  |  |  |
| --- | --- | --- | --- | --- |
|  | 1st | 2nd | 3rd | Total |
| First | 11 (73.3) | 12 (80.0) | 10 (66.7) | 14 (93.3) |
| Second | 12 (80.0) | 8 (53.3) | 11 (73.3) | 15 (100) |

**Figure 8.**
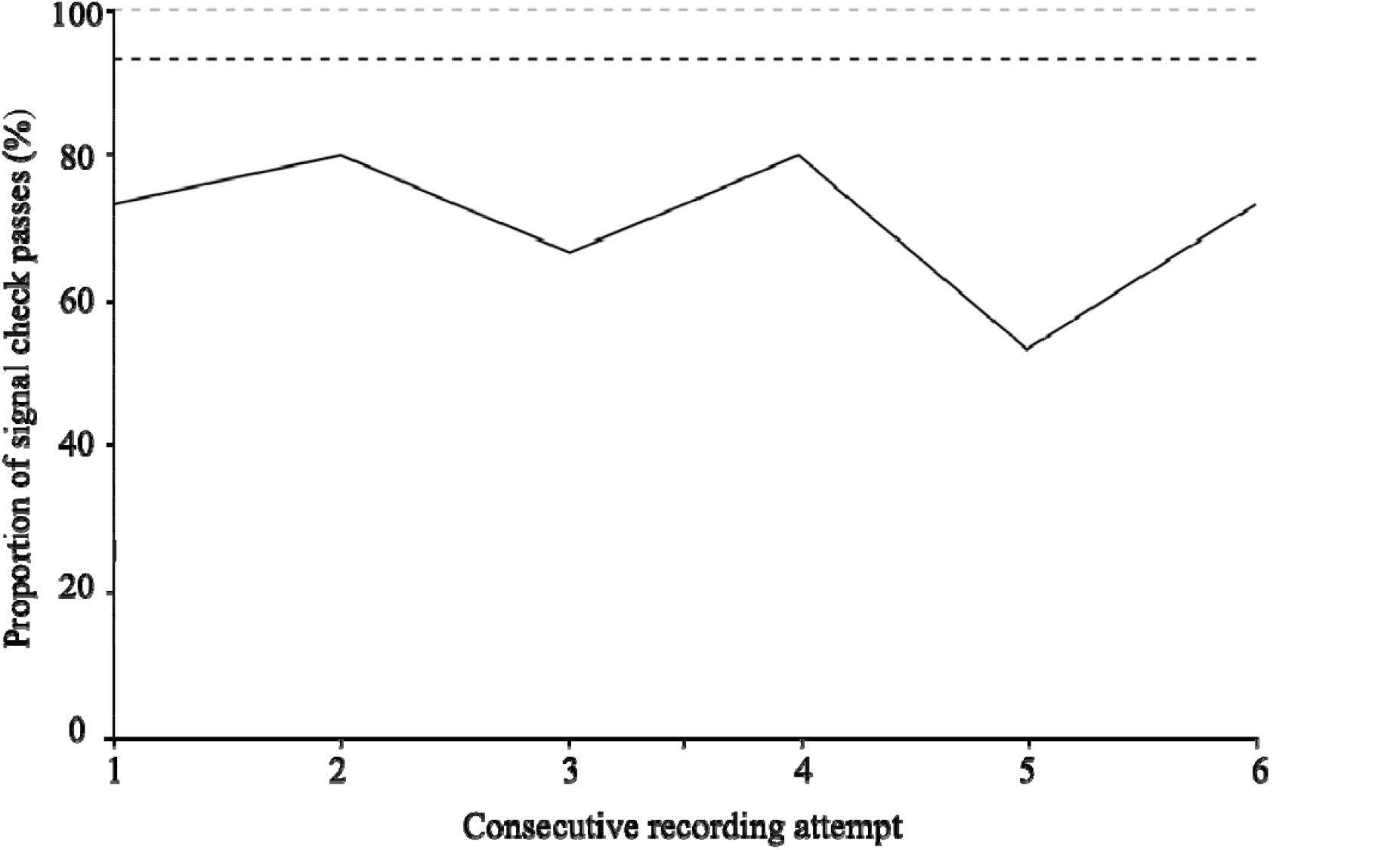
Line graph indicating the proportion of Study 1 participants who were able to generate a recording that passed the signal check on each of six consecutive recording attempts. The black dashed line indicates the proportion of individuals who were able to generate a recording that passed the signal check by the end of their first use of the system (three consecutive recordings), and the grey dashed line indicates the proportion of individuals who were able to do so by the end of their second use of the system.

#### Interim conclusion

Study 1 results indicated strong relationships between the FDA-cleared reference and both the mHealth app and the novel reference. Notably, these relationships were highly comparable to functional outcomes for alternative FDA-cleared RR monitoring devices.^12, 14, 36, 38^ Accordingly, these results supported both continued assessment of the mHealth app and application of the novel reference to accuracy analyses in Study 2, as described below.

## Study 2

### Methods

Study 2 aimed to establish the accuracy of the mHealth app ‘in the wild’ via remote data capture, compared to the novel reference validated in the Study 1. The usability of the mHealth app was additionally assessed in a larger sample. Measures and statistics were as described for Study 1.

#### Participants and recruitment

Participants were recruited via an online research platform, with study enrollment controlled to ensure a proportionate distribution of age, gender and smartphone ownership (iOS versus Android). Inclusion criteria included being aged 18 or over, having access to a smartphone of minimum requirements to download the mHealth app and being willing and able to follow the study protocol and complete an informed consent form. As researchers would not monitor participants during their participation, additional safety criteria excluded individuals who were pregnant, breastfeeding, had a pacemaker, or a chest or spine problem that could affect their breathing.

#### Procedure

Participants were directed to online documentation containing full information about the study procedures before completing an online eConsent procedure. They then completed a baseline questionnaire concerning their demographics, including age, sex, ethnicity, height and weight, before receiving instructions to download and activate the mHealth app.

Participants were requested to follow instructions provided within the mHealth app to capture 10 RR recordings, including recordings that both passed and failed the signal check, before completing a System Usability Scale (SUS)^39^ and providing separate qualitative feedback on their experience using the mHealth app. Study-specific procedures took approximately 20 minutes, for which participants were reimbursed £2.50 through the research platform.

## Results

### Participants and data

165 participants enrolled in the study, of whom 152 completed the baseline questionnaire concerning their demographics (Table 2). Medical conditions reported included asthma (respiratory), arthritis and Parkinson’s disease (movement). 5 participants were excluded due to significant deviation from the study protocol, resulting in a participant cohort of 160, for whom a mode of 11 mHealth app recordings each was captured. 987 recordings passed the signal check and were included in accuracy analyses. Recordings were submitted from 64 unique smartphone models, 46 (71.9%) of which were Android and the rest were iPhone models.

**Table 2.** Demographic characteristics for Study 2 participants (n = 152).

| Characteristic | Female (n = 67) | Male (n = 85) | Total (n = 152) |
| --- | --- | --- | --- |
| Age in years, mean (SD, range) | 43.5 (14.85, 18-73) | 40.7 (14.70, 19-69) | 41.9 (14.78, 18-73) |
| Weight in kg, mean (SD, range) | 70.2 (19.45, 47-159) | 87.9 (22.31, 52-203) | 80.1 (22.08, 47-203) |
| Height in m, mean (SD, range) | 1.66 (0.108, 1.45-2.16) | 1.78 (0.660, 1.56-1.93) | 1.73 (1.061, 1.45-2.16) |
| BMI in kg/m<sup>2</sup>, mean (SD, range) | 25.4 (6.28, 17.8-58.4) | 27.7 (6.80, 17.4-62.5) | 26.7 (6.65, 17.4-62.7) |
| **Ethnicity, n (%)** |  |  |  |
| White | 57 (85.1) | 72 (84.7) | 129 (84.9) |
| Asian | 4 (6.0) | 7 (8.2) | 11 (7.2) |
| Black | 3 (4.5) | 2 (2.4) | 5 (3.3) |
| Mixed/multiple | 3 (4.5) | 4 (4.7) | 7 (4.6) |
| **Medical conditions, n (%)** |  |  |  |
| Respiratory disorder | 9 (13.4) | 7 (8.2) | 16 (10.5) |
| Movement disorder | 2 (3.0) | 0 (0) | 2 (1.3) |
| Cognitive disorder | 0 (0) | 0 (0) | 0 (0) |

### Accuracy

Error results indicated an MAE of 1.14 BPM (SD = 1.44) with a 95% CI of 1.02 - 1.26. Relative MAE was 9.5% (SD = 18.70) with 95% CI of 8.38 - 10.72. Bias (novel reference – mHealth app) was 0.08 (SD = 1.84) with LoA of -3.68 - 3.51, indicating slight RR underestimation by the mHealth app. 61 comparisons (6.2%) had an absolute error greater than 3 BPM. A Bland-Altman plot indicated error values as a function of RR averaged between the reference and mHealth app (Figure 9). PPMC produced a coefficient of r(986) = 0.855, p < .000, indicating a high or strong association^37^ (Figure 10).

**Figure 9.**
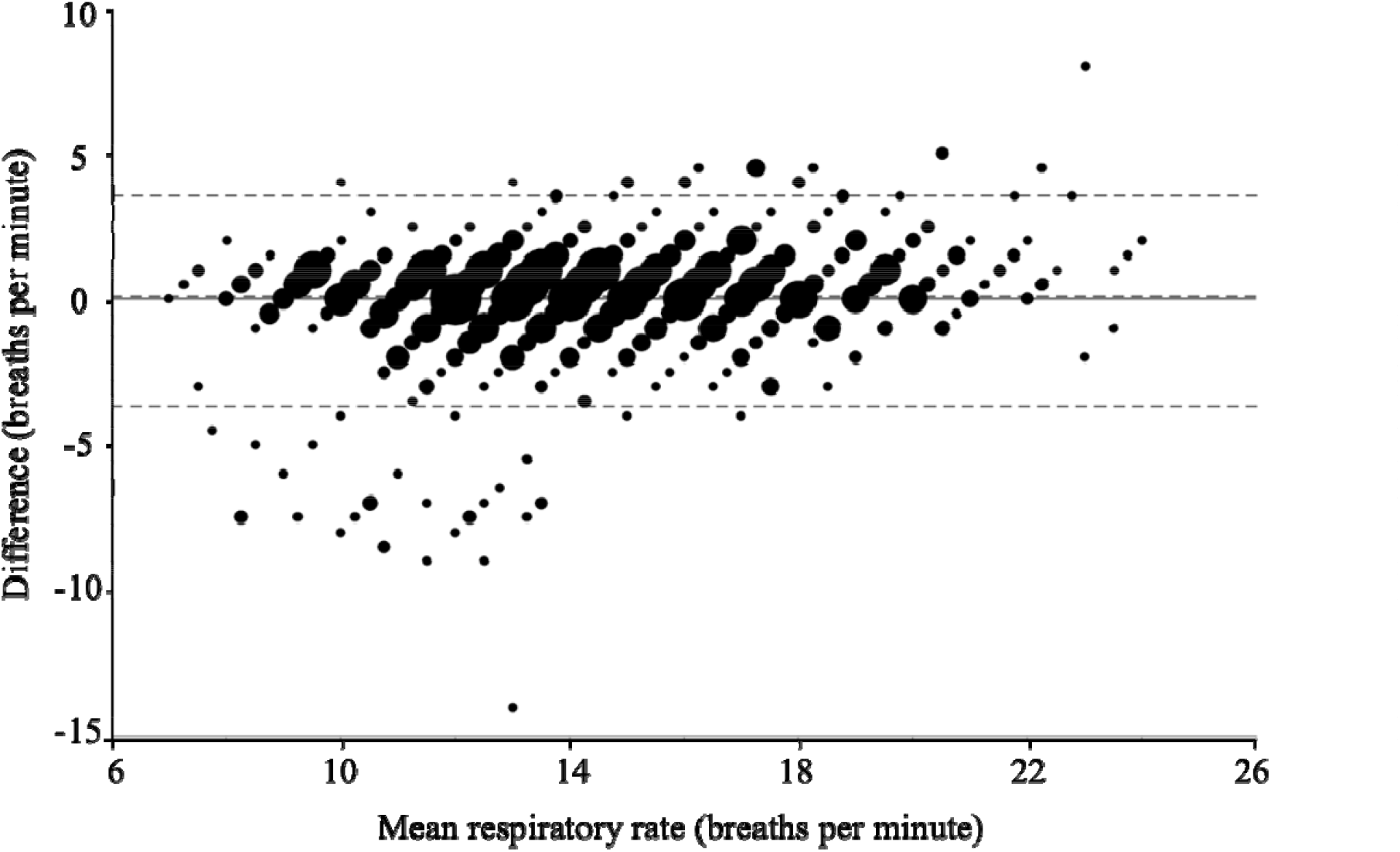
Bland-Altman plot for RR estimates provided by the mHealth app and novel reference. The x-axis indicates RR estimates averaged between the mHealth app and novel reference and the y-axis indicates the difference between RR estimates from each source (novel reference - mHealth app). The solid horizontal line depicts a mean difference (bias) of 0 and dashed lines from top to bottom represent the upper limit of agreement (3.51), the observed mean difference (bias; 0.08), and the lower limit of agreement (−3.68). Marker size is proportional to the number of observations for each combination of values.

**Figure 10.**
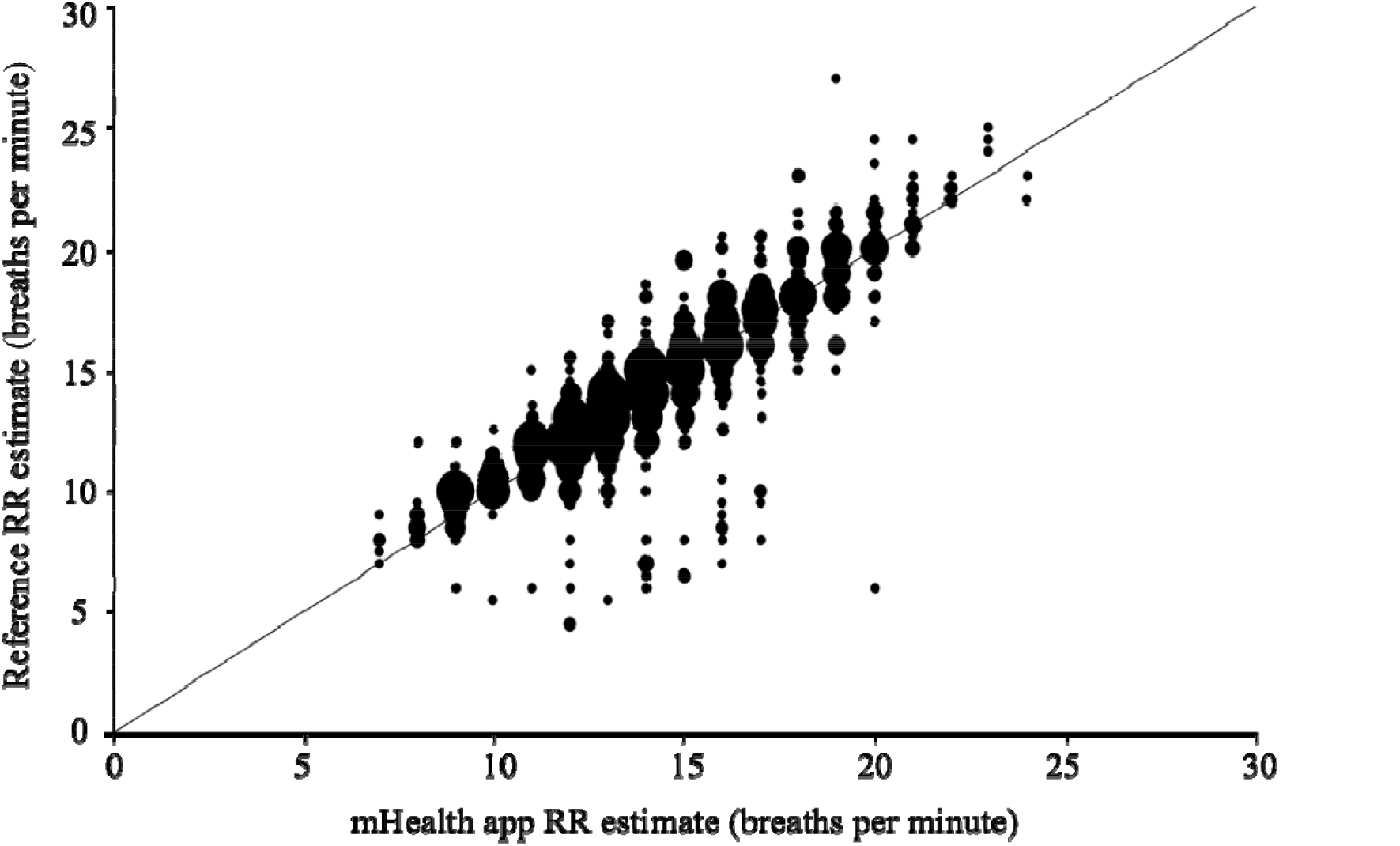
Scatterplot for simultaneous RR estimates provided by the mHealth app (x-axis) and novel reference (y-axis). The solid line indicates the gradient y=x. Marker size is proportional to the number of observations for each combination of values.

### Usability

149 (93.1%) of a total of 160 participants who captured mHealth app recordings were able to use the system successfully on their first try (Table 3; Figure 11). 155 (96.9%) did so by their second try.

**Table 3.**
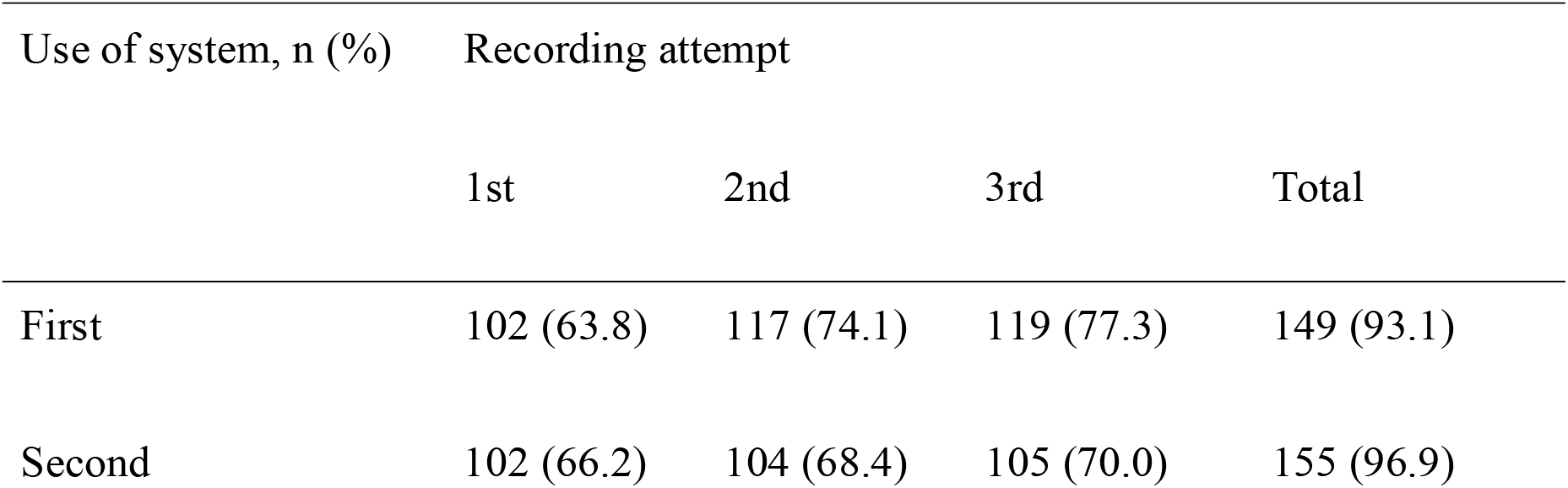
Number and proportion of participants able to use the system successfully on consecutive attempts in Study 2 (n = 160).

**Figure 11.**
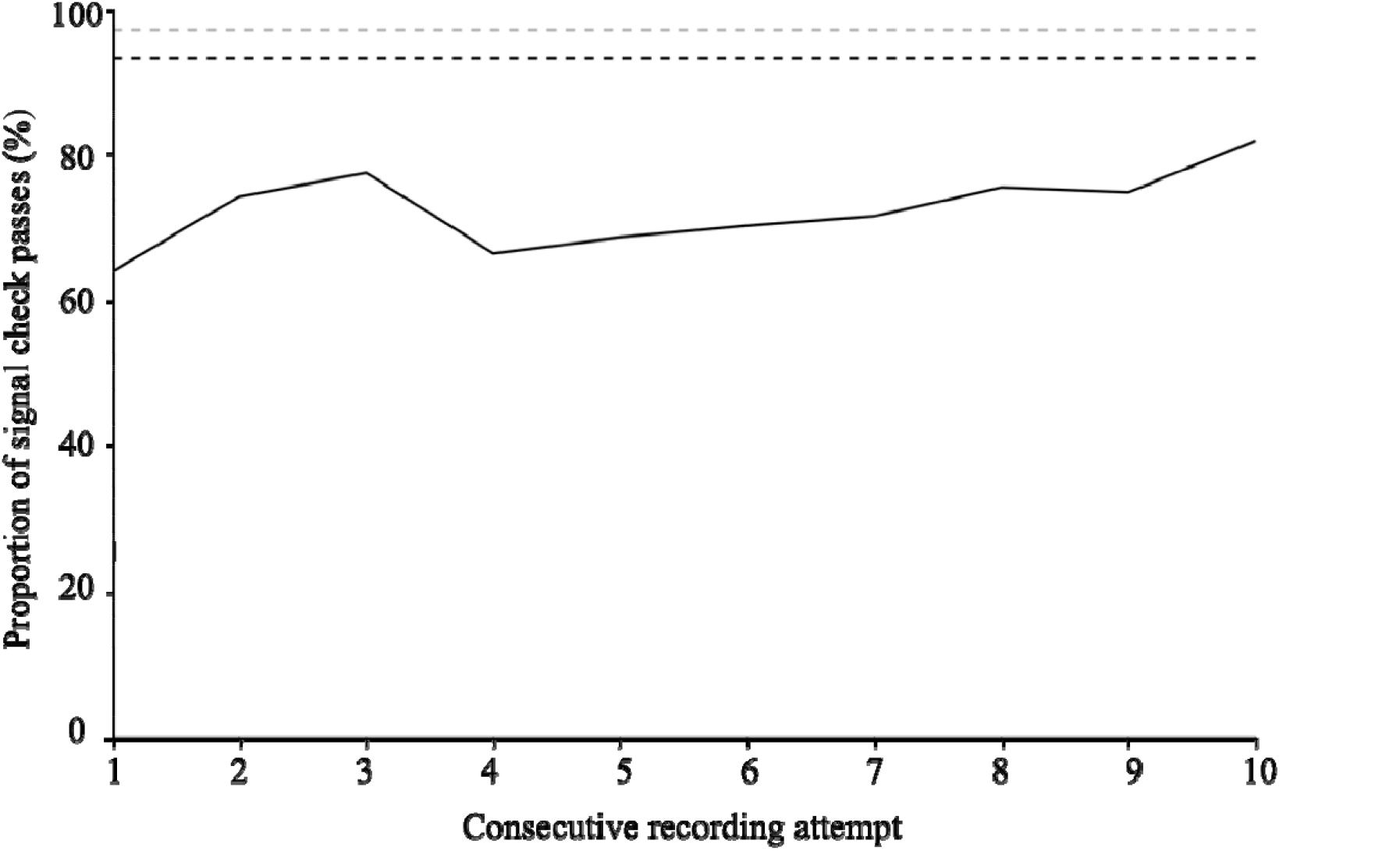
Line graph indicating the proportion of Study 2 participants who were able to generate a recording that passed the signal check on each of ten consecutive recording attempts. The black dashed line indicates the proportion of individuals who were able to generate a recording that passed the signal check by the end of their first use of the system (three consecutive recordings), and the grey dashed line indicates the proportion of individuals who were able to do so by the end of their second use of the system.

The mean SUS score was 73.2 (SD = 5.39). Of the sub-scales, each scored between 0 and 4, those most agreed with by participants were: *I would imagine that most people would learn to use this system very quickly* (3.2), *I thought the system was easy to use* (3.1) and *I felt very confident using the system* (3.0). The lowest scoring, indicating participant disagreement, were: *I think that I would need the support of a technical person to be able to use this system* (0.5) and *I needed to learn a lot of things before I could get going with this system* (0.8).

## Discussion

### Principal findings

To the authors’ knowledge, this is the first study to assess at scale a user-operated novel mHealth smartphone application designed to capture a user’s RR using smartphone movement sensors, considering both accuracy and usability in an ecologically valid study environment. Outcomes for the mHealth app were highly comparable to results published for medical devices available on the market today (Table 4). In addition, as changes in breathing rate greater than 3 BPM may indicate clinical deterioration^35-36^, observations that error values for the mHealth app were typically less than this threshold suggest the device may carry low clinical risk. Study 2 revealed a small cluster of substantial overestimation errors (5-10 BPM) for lower RRs (8-14 BPM). Although this observation was not found in the Study 1, this may be due to the smaller sample size in that analysis. The nature of these overestimations is unclear based on the present analyses. Overestimation of RR carries clinical risk with regard to both underdiagnosis of bradypnea (low RR) and overdiagnosis of tachypnea (elevated RR) that may lead to clinical decision making based on misinformation, although it should be noted that RR is rarely used in isolation to inform clinical decision making. Future research should seek to identify and mitigate the cause of these errors.

**Table 4.** Comparison of mHealth app results to alternative devices available on the market today.

| Comparison | MAE (SD, 95% CI) | Relative MAE (SD, 95% CI) | Bias (SD, LoA) | Correlation coefficient |
| --- | --- | --- | --- | --- |
| **mHealth app** |  |  |  |  |
| Compared to FDA-cleared reference | 1.65 (1.49, 1.32 - 2.06) | 12.2 (9.23, 10.06 - 14.57) | 0.81 (2.08, - 3.27 - 4.89) | 0.700 |
| Compared to novel reference | 1.14 (1.44, 1.02 - 1.26) | 9.5 (18.70, 8.38 - 10.72) | 0.08 (1.84, - 3.68 - 3.51) | 0.885 |
| **Respirasense (PDM Solutions)** |  |  |  |  |
| Compared to capnography<sup>12</sup> | - | - | 0.38 (N/A, N/A, -1.0 - 1.8) | - |
| Compared to manual count<sup>12</sup> | - | - | -0.70 (N/A, N/A, -4.9 - 3.5) | - |
| Compared to electrocardiogram<sup>36</sup> | - | - | -0.41 (1.79, - 0.73 - -0.08, - 3.9 - 3.1) | 0.84 |
| Compared to manual count<sup>36</sup> | - | - | -0.58 (2.5, - 1.04 - -0.12, - 5.5 - 4.3) | 0.78 |
| BioStamp nPoint (MC10) compared to capnography<sup>14</sup> | 1.3 (2.1, N/A) | - | -0.29 (N/A, N/A, -5.17 - 4.59) | 0.697 |
| Rad-87 (Masimo) compared to capnography<sup>38</sup> | - | 10 (9, 7-13) | - | - |

Concerning usability, most participants could successfully operate the mHealth app on their first or second use of the system. Although no industry standards for successful operation exist, results observed here appeared to be broadly similar to values that could be estimated from available literature regarding other physiological measurement mHealth apps, which were typically in the range of 95% or higher.^40-42^ Subjective usability outcomes were also promising, with an overall SUS score well above the industry average of 68.^43^ Study 2 revealed a general trend of high signal check pass rates for later sequential recording attempts, suggesting that participants found it easier to capture RR recordings the more they used the mHealth app. Although this learning effect was not observed in the Study 1 results, this may be due to observer bias and a small sample size within that study setting. This observation holds promise for improved usability with long-term use of the mHealth app, although it may indicate greater clinical risk during early use of the system. Future research may seek to steepen the learning curve to minimise clinical risk.

Strengths of the present study include the application of a remote study design that lends ecological validity to the results and selective recruitment to ensure a heterogeneous participant cohort, which suggests good generalisability of the results. In addition, the inclusion of usability assessment allows a holistic perspective on the mHealth app to be generated. In all, these results hold promise for the use of smartphone movement sensors as a viable means of remote RR monitoring. Software-based mHealth may offer cost and scalability benefits compared to hardware-based monitoring. Additionally, movement sensors may better protect RR signal quality than alternative devices that use microphone and camera sensors, as these are vulnerable to noise from environmental light and sound that is difficult to control. These benefits suggest that RR monitoring based on smartphone movement sensors may support healthcare systems to care for their patients when they are outside of the clinic better than currently available alternatives.

## Study limitations

As only healthy participants were recruited, it is also unclear how the observed results may extrapolate to healthcare patients who would be likely real-world users of the mHealth app - particularly those with abnormal breathing rates and patterns due to a respiratory condition. Participants both from Study 1 (employees of the mHealth app manufacturer) and Study 2 (members of an online research community) were likely to be technologically confident and may have therefore been predisposed to successfully operating the mHealth app. Future research should seek to incorporate individuals of low technical literacy and target end-users with relevant medical conditions to better understand these results’ generalisability.

Concerning methodology, the FDA-cleared reference used in Study 1 has its own measurement error.^13^ Hence, error estimates presented here are, in fact, an unknown combination of errors associated with the FDA-cleared reference and mHealth app versus true RR. The Study 2 reference also underwent only limited validation in Study 1 and should be assessed more rigorously. Future research may wish to apply a wider range of reference methods, including gold and industry-standard references, to reduce the vulnerability of the mHealth app to shortcomings of any single reference.

Additionally, the present research design does not directly address potential benefits the mHealth app may offer if applied in a healthcare setting. Although expectations that moving health assessments outside of a clinical setting via mHealth technologies will improve healthcare economics have been somewhat supported by literature,^4^ clinical evidence suggests that mHealth technologies are highly heterogeneous in their ability to improve health outcomes.^44-45^ Suggestions that mHealth may help to overcome social, economic and geographical barriers to healthcare are also yet to be validated.^46-48^ Future research should seek to understand the clinical, economic and social outcomes associated with real-world use of the mHealth app.

## Conclusions

Decentralised healthcare technology holds the potential to offer clinical and economic benefits to patients, HCPs and healthcare systems. Breathing is an important indicator of health, and although solutions for remote RR monitoring exist, many entail significant shortcomings that may limit their ability to capitalise on potential benefits of mHealth. Results from the present study hold promise for the use of smartphone movement sensors as a robust means for remote RR monitoring. However, future research should address residual questions and risks associated with the technology identified in this article and seek to validate the impact of similar technologies as applied in the real world.

## Data Availability

All data produced in the present study are available upon reasonable request to the authors

## Acknowledgements

The authors are grateful to Huma’s wonderful developers Michele Colombo, Stanislas Heili, Leonardo Festa, Davide Mascitti, Emanuele Distefano, Matteo Puccinelli and Matteo Vigoni for their diligent efforts in preparing the mHealth app for this research. Additional thanks to Emily Binning for her continued strategic support that helped this research come to fruition.

## Funding

This research was sponsored by Huma Therapeutics Ltd.

## Ethical Approval

Ethical approval was provided by the University of Exeter’s Research Ethics Board (application ID eUEBS004088) and all research was conducted in compliance with the Declaration of Helsinki.

## Guarantor

DP accepts the responsibility of acting as Guarantor.

## Conflicts of Interest

All authors are current or previous employees of Huma Therapeutics Ltd, which is the developer of the mHealth smartphone app. No additional conflicts of interest relevant to this study are declared.

## Author Contributions

SP, BK, MD, and MB acquired, interpreted and aided in drafting and revising of the article for intellectual content. DP aided in data interpretation and drafting and revising of the article for intellectual content.

### Abbreviations

apps: applications
BPM: breaths per minute
FDA: US Food and Drug Administration
HCP: healthcare professional
LoA: Limits of agreement
MAE: mean absolute error mHealth mobile health
PPMC: Pearson Product Moment Correlation
RR: respiratory rate
SD: standard deviation
SUS: System Usability Scale

## Notes

**Conflict of Interest:** SV, BK, MD, MB, DP, are or were employees of Huma Therapeutics Ltd at the time of writing.

### Funding Statement

This study was funded by Huma Therapeutics Ltd.

